# Antimalarial Drug Resistance Profiling of *Plasmodium falciparum* Infections in India Using Next-Generation Sequencing

**DOI:** 10.1101/2023.04.08.23288321

**Authors:** Sonal Kale, Swapna M. Uplekar, Nabamita Bandyopadhyay, Pavitra N. Rao, Syed Z. Ali, S.K. Sharma, Nikunj Tandel, Ankita Patel, Ranvir Singh, Aaron Dank, Sangamithra Ravishankaran, G Sri Lakshmi Priya, Aswin Asokan, Alex Eapen, Om. P. Singh, Jane M. Carlton, Prashant K. Mallick

**Affiliations:** National Institute of Malaria Research, Indian Council of Medical Research, Sector 8, Dwarka, New Delhi, India; Laboratory of Malaria Immunology and Vaccinology, National Institute of Allergy and Infectious Disease, NIH, Bethesda, Maryland, USA; Center for Genomics and Systems Biology, Department of Biology, New York University, New York, NY, USA; National Institute of Malaria Research Field Unit, Sector 1 Health Center, Rourkela, Odisha, India; National Institute of Malaria Research Field Unit, Civil Hospital, Nadiad, Gujarat, India; National Institute of Malaria Research Field Unit, Indian Council of Medical Research, National Institute of Epidemiology Campus, Ayapakkam, Chennai, Tamil Nadu, India

**Keywords:** India, *Plasmodium falciparum*, antimalarial drug resistance, deep sequencing, genomic

## Abstract

**Background:** Tracking the emergence and spread of antimalarial drug resistance has become critical to sustaining progress towards the control and eventual elimination of malaria in South Asia, especially India.

**Methods:** An amplicon sequencing protocol was used for high-throughput molecular surveillance of antimalarial drug resistance in a total of 158 isolates at three sites in India: Chennai, Nadiad and Rourkela. Five genes of the *Plasmodium falciparum* implicated in antimalarial resistance were investigated here; *Pfcrt* for chloroquine resistance, *Pfdhfr* for pyrimethamine resistance, *Pfdhps* for sulfadoxine resistance, *Pfk13* for artemisinin resistance and *Pfmdr1* for resistance to multiple antimalarials.

**Results:** Mutations in the propeller domain of PfK13 were observed in two samples only, however these mutations are not validated for artemisinin resistance. A high proportion of parasites from the *P. falciparum* dominant site Rourkela showed wild-type *Pfcrt* and *Pfdhfr* haplotypes, while mutant *Pfcrt* and *Pfdhfr* haplotypes were fixed at the *P. vivax* dominant sites Chennai and Nadiad. The wild-type PfDHPS haplotype was predominant across all study sites. Finally, we observed the largest proportion of suspected multi-clonal infections at Rourkela, which has the highest transmission of *P. falciparum* among our study sites.

**Conclusion:** This is the first simultaneous high-throughput next generation sequencing of five complete *P. falciparum* genes from infected patients in India.

## Background

Malaria remains a major cause of mortality and morbidity in tropical and sub-tropical countries. In 2015, Africa had the greatest malaria burden with 88% of global malaria cases, followed by Southeast Asia with 10%, and the eastern Mediterranean region with 2%[1]. India reported 1.2 million microscopy-confirmed cases of malaria and 384 deaths due to malaria in 2015, accounting for approximately 89% of malaria cases in South East Asia and 6% of the total malaria cases in the world [2]. Over 60% cases in India are caused by *Plasmodium falciparum*. Massive efforts by a worldwide malaria elimination program have reduced the number of malaria cases from 2010 by 12% to 214 million in year 2015, and reduced the number of malaria deaths by 21% [3]. India has also joined malaria elimination efforts, allocating financial resources and infrastructure for the launch of a ‘National Framework for Malaria Elimination in India 2016-2030’ [1, 4]. This effort includes all Indian states with varying levels of malaria transmission and outlines specific strategies to achieve the following milestones: 1) eliminate malaria throughout the entire country by 2030; and 2) maintain a malaria-free status in areas where transmission has been interrupted and prevent re-introduction of malaria.

The emergence and spread of parasite resistance against available antimalarials constitutes a major threat towards the efforts of the elimination program, as evidenced by the reversal of previous malaria eradication successes to failure [5]. Routine molecular surveillance of parasite strains is an essential tool for identifying and limiting the spread of emerging antimalarial resistance. Sanger sequencing is the gold standard for validation of genotypes identified in parasite clinical isolates, but sensitivity and scalability remain limiting factors; these may be overcome by high-throughput next generation sequencing (NGS) that facilitates the generation of massive amounts of genomic data in a relatively short time by parallelization of sequencing reactions. The technology provides high sequencing coverage, which enables the detection of low-frequency mutations relevant to drug resistance, or low frequency wild-type parasites arising after reversion of mutant genotypes following antimalarial withdrawal from a region. NGS can also provide deep insight into genetic diversity of heterogeneous parasite populations.

We have previously described a novel amplicon sequencing protocol for high throughput molecular surveillance of drug resistance in field settings [6]. This protocol identifies polymorphisms in five important genes implicated in *P. falciparum* resistance against the mainstay antimalarials; *Pfcrt* for chloroquine resistance, *Pfdhfr* for pyrimethamine resistance, *Pfdhps* for sulfadoxine resistance, *Pfk13* for artemisinin resistance and *Pfmdr1* for resistance to multiple antimalarials. The Center for the Study of Complex Malaria in India (CSCMI) was established in 2010, with the aim of elucidating the epidemiology and transmission of malaria in India at three ecologically diverse sites - Chennai, Nadiad and Rourkela [7]. In this study, we have implemented the aforementioned amplicon-sequencing protocol in field settings to elucidate genomic landscapes of drug resistance at these three CSCMI study sites representing varying transmission ecologies. To our knowledge, this is the first study providing status of multiple drug resistance genotypes at these sites.

## METHODS

### Malaria epidemiology of study sites

Chennai, located on the eastern coast of India is the capital city of Tamil Nadu state, and had over 7 million inhabitants in 2011. Chennai accounts for 55.6% of all malaria cases in Tamil Nadu, which had an annual parasite index (API) of 0.12 in 2014 [4, 8]. Malaria is hypo-endemic in Chennai, and *P. vivax* is the dominant species. Subjects were enrolled at the Besant Nagar Malaria Clinic, or in cross-sectional surveys conducted in the Besant Nagar catchment area. Nadiad, located in the Kheda district in Gujarat state (API in 2014: 0.66), had a population of 0.2 million in 2011. Nadiad is hypo-endemic for malaria, with slightly higher prevalence of *P. vivax* than *P. falciparum*. Subjects were enrolled at either the NIMR malaria clinic in Nadiad Civil Hospital or at an urban health center in the Vatva ward of Ahmedabad city, the capital of Gujarat state. Alternately, subjects were enrolled in cross-sectional surveys conducted in the vicinity of Nadiad town. Rourkela, located in Sundargarh district close to the northern border of the state of Odisha (API in 2014: 9.08), had a population of over 0.5 million in 2011. Rourkela has meso-to-hyperendemic transmission of malaria, with *P. falciparum* as the dominant species. Subjects were enrolled at a health center clinic set up in a suburb of Rourkela, and in cross-sectional surveys conducted in rural areas around Rourkela.

### Sample collection and processing

158 of 300 *P. falciparum* isolates collected as part of the CSCMi study [7] and tested using our amplicon sequencing protocol [6] were sequenced successfully, with amplification failing in isolates with poor quality or low concentration of DNA. These isolates were collected from 141 study enrollees at three sites: Chennai (18 isolates from 15 patients), Nadiad (55 isolates from 41 patients) and Rourkela (85 isolates from 85 patients), between December 2012 and September 2015 (**Table 1**). A majority of the enrollees were male (98 male, 43 female). Patients were between 2-69 years old, with a mean age of 27.44 (95%CI: 24.83-30.07). Asexual or sexual *P. falciparum* parasitemia was detectable by microscopy in 140 isolates, with a mean parasitemia of 14073.16 parasites/ul (95%CI: 3822.15-24324.18); 18 isolates were submicroscopic. 149 isolates were positive for *Plasmodium falciparum* species-specific PCR, 9 were PCR negative. Patients were enrolled after obtaining informed consent, and treated with antimalarials as per the Indian national guidelines [9].

**Table 1:**
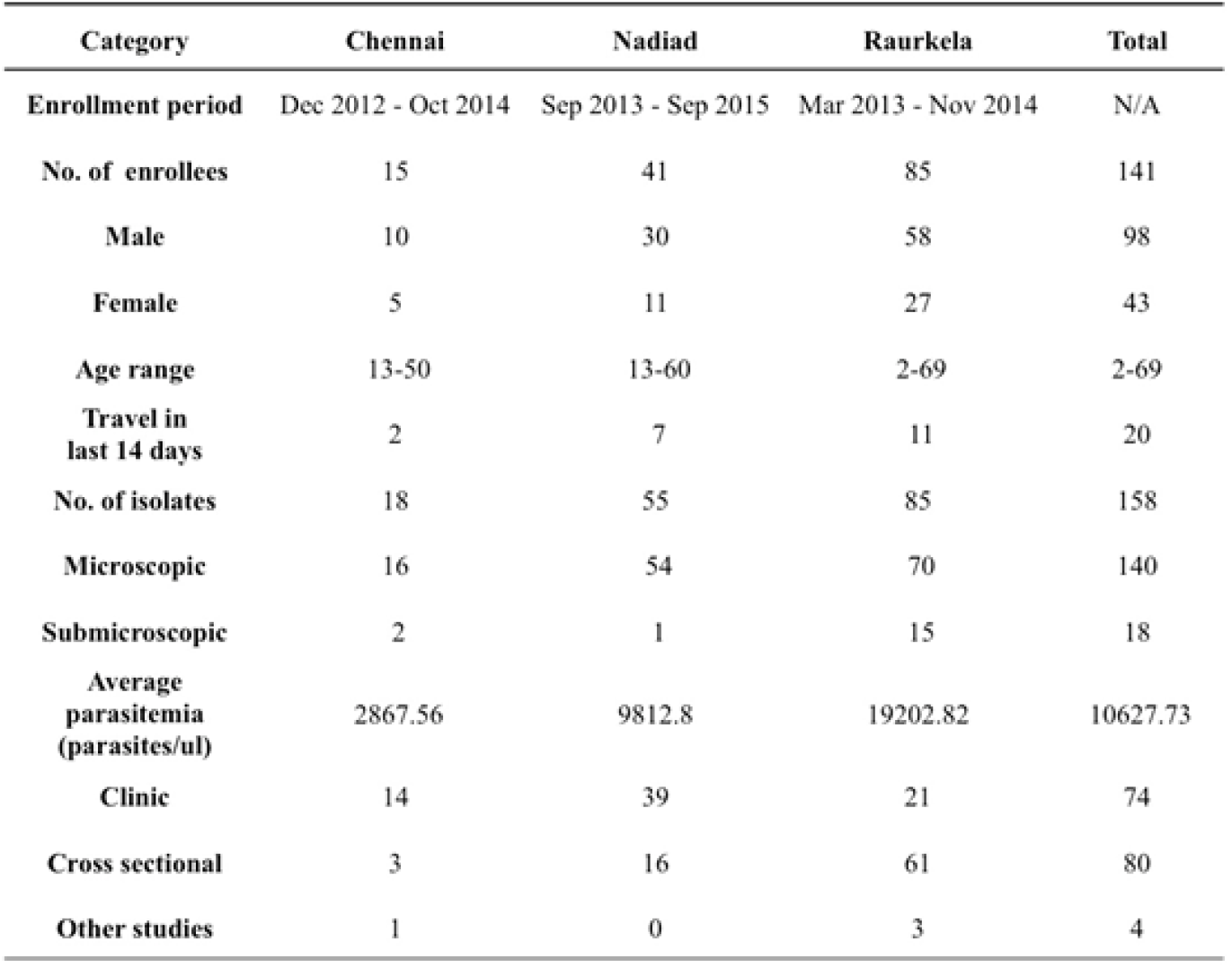
Sample summary and descriptive statistics

### DNA extraction, library preparation and sequencing

DNA was extracted from *P. falciparum-*infected blood samples collected on Whatman filter paper, small EDTA microvettes or EDTA vacutainers using QIAamp DNA Blood Mini or Midi Kit (Qiagen Inc., catalog nos. 51106, 51185) after separation of plasma, and 1-5 ul was used as a template for amplicon synthesis. Primers and PCR conditions were as outlined in [6]. For each sample, 10 ul of every amplicon was pooled, and the pooled mixture was purified using the QIAquick PCR Purification Kit (Qiagen Inc., catalog no. 28106) and quantitated using a Qubit 2.0 fluorometer (Thermo Fisher Scientific, catalog no. Q32866). Barcoded sequencing libraries were generated from 100-500 ng of purified pooled PCR product using ion express™ barcode adapters (Life Technologies, catalog no. 4471250 for barcodes 1-16; catalog no. 4474009 for barcodes 17-32) and library prep kit (New England Biolabs, catalog no. E6285L). The size and quality of libraries were determined using an Agilent bioanalyzer, and quantified by quantitative PCR using Ion Torrent Library Quantification kit (KK4857, KAPA biosystems) on a LightCycler^R^ 480 (Roche). For most experiments, 16 barcoded libraries were pooled for a single run on the PGM using Ion PGM™ Hi-Q™ sequencing technology and Ion™ 314v2 type chips, except for one library pool that was run on an Ion™ 318v2 chip. Genomic DNA from *P. falciparum* strains Dd2 (MRA-150G) and 7G8 (MRA-152G) was obtained from the Malaria Research and Reference Reagents Resource (MR4). Control libraries prepared with Dd2 or 7G8 DNA were included in each run. We also sequenced ten additional *P. falciparum* reference strains (NF54 E, TM91C235, HB3, W2, K1, V1/S, D10, GB4, D6, FCB) as part of different runs to ensure that our variant calling results showed complete concordance with previously published data for these isolates.

### Data processing

Sequencing data were analyzed using the Ion Torrent platform software (Torrent Suite 5.0.2). Raw reads were de-multiplexed and filtered using standard quality filtering parameters by Torrent Suite pipeline software. Read quality was assessed using the Torrent Suite FastQC plugin v0.10.1, and high-quality reads were aligned to the reference using the Torrent Mapping Alignment Program v5.0. The reference data file used was a multi-FASTA file containing the complete gene sequence and 300bp flanking regions for each amplicon target gene from the P. falciparum 3D7 genome using the PlasmoDB database.

### Data analysis

Variant calling was performed with low stringency parameters using the Ion Torrent Variant Caller plugin (v5.0). We generated a list of all positions in all genes that were variable in at least one sample; for samples where no variants were reported at these positions, the presence of the reference allele was confirmed by obtaining raw nucleotide counts and quality scores from corresponding BAM files using BEDTools. Variant filtering criteria was determined based on the quality of known SNPs in MR4 reference isolates sequenced in each run. Variant calls with SNP allele depth < 10 reads and base quality score < 10 were discarded. Calls with SNP allele frequency between 20% - 80% were marked as heterozygous and subsequently used to classify samples as multi-clonal infections, i.e. containing more than one distinct parasite genotype. SNP calls were processed to determine codon changes based on the *P. falciparum* 3D7 gene annotation from the PlasmoDB database using custom Perl and MS Excel VBA scripts. In the case of multiple nucleotide polymorphisms (MNPs) affecting the same codon, if both SNP alleles occur at a frequency > 80%, they were merged to calculate the resulting codon change; variant calls with SNP allele frequency < 20% were discarded. For the *Pfcrt* gene, MNPs affecting codon positions 72-76 were filtered using a higher SNP allele frequency cut-off of 30%. A high proportion of homo-polymer repeats in the *Pfcrt* gene led to poor sequencing quality misalignment of reads. As a result variants in this gene were manually curated using the IGV genome browser, particularly for the genomic locus corresponding to codons 72-76. Population structure was examined using a non-model-based multivariate analysis method as implemented in the adegenet 2.0.1 package using the *dudi*.*pca* function. In the case of multi-allelic SNPs, the major allele call was used to represent the dominant genotype at that position.

## RESULTS

We have successfully sequenced a panel of five *P. falciparum* genes implicated in antimalarial drug resistance in clinical isolates from India using a previous validated amplicon sequencing protocol [6]. *Pfk13, Pfcrt, Pfmdr1, Pfdhfr* and *Pfdhps* have been implicated in resistance to first-line antimalarial therapy, including artemisinin combination therapy, chloroquine, and sulfadoxine-pyrimethamine. We deduced SNP haplotypes from key resistance-associated alleles in the target genes for 158 isolates collected at three sites in India. Implications of the resistance mutations identified at the three sites have been described below in the context of each drug they are associated with.

### Resistance to chloroquine

Mutations encoding amino acid substitutions in the chloroquine resistance transporter PfCRT (PF3D7_0709000) at positions 72, 74, 75, 76, 97, 220, 271, 326, 356 and 371 have been associated with increased resistance to chloroquine (CQ), with the K76T substitution being the primary mediator of resistance [10-12]. 147 of 158 isolates sequenced carried only one haplotype at the PfCRT locus; one isolate had multiple PfCRT haplotypes, and 10 isolates had partial or missing PfCRT haplotype data (**Table 2**). The CQ-resistant PfCRT haplotype S^72^V^73^M^74^N^75^T^76^, purported to be the most prevalent PfCRT haplotype in India [13], was present in 71/147 isolates, including all isolates from Chennai and most isolates from Nadiad, but only two isolates from Rourkela. Contrastingly, the CQ-sensitive wild-type haplotype C^72^V^73^M^74^N^75^K^76^ was detected in 46 isolates from Rourkela, but absent from Nadiad and Chennai. The CQ-resistant haplotype C^72^V^73^I^74^E^75^T^76^ was present in 30/147 isolates: 28 isolates from Rourkela and 2 isolates from Nadiad. Both copy number variation and mutations encoding substitutions in the multidrug resistance protein PfMDR1 (PF3D7_0523000) codons 86, 184, 1034, 1042 and 1246 can modulate resistance to various antimalarial drugs (summarized in [14]). While our protocol does not permit gene copy number assessment, we were able to identify polymorphisms associated with resistance to CQ. 129 of 158 isolates sequenced carried only one PfMDR1 haplotype, while 10 isolates possessed more than one PfMDR1 haplotype, which may be attributed to increased *Pfmdr1* copy number (**Table 2**). 19 isolates had partial or missing haplotype data. The single-mutant PfMDR1 haplotype N^86^F^184^S^1034^N^1042^D^1246^ was predominant (57/129) among isolates in our study, and the wild-type PfMDR1 haplotype N^86^Y^184^S^1034^N^1042^D^1246^ was also prevalent in several (45/129) isolates. Double-mutant PfMDR1 haplotypes were rare, with only 5/129 isolates containing the haplotypes Y^86^F^184^S^1034^N^1042^D^1246^ or N^86^F^184^S^1034^D^1042^D^1246^. The single mutation-bearing haplotype Y^86^Y^184^S^1034^N^1042^D^1246^ was present in 22/129 isolates, 11 each from Nadiad and Rourkela.

**Table 2:**
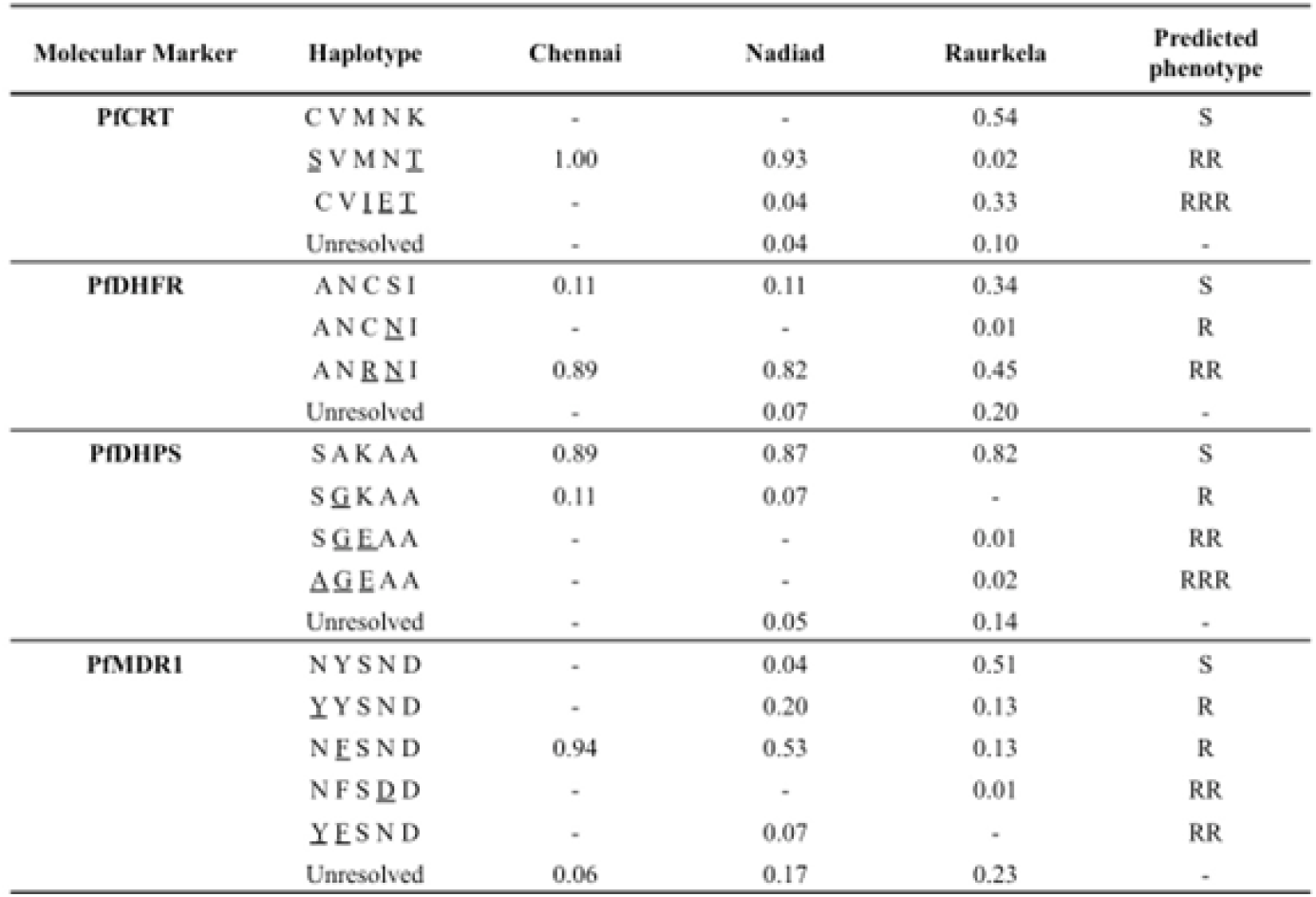
Frequency of haplotypes associated with drug resistance at three sites in India

A total of 12 haplotype combinations between the PfCRT and PfMDR1 loci were seen in 122 isolates with complete sequence information for these genes across the three sites. Rourkela had the largest diversity of haplotypes, with 10 haplotype combinations at these two loci, and Chennai had the least diversity (**Table 3**), with only one haplotype combination. The PfCRT and PfMDR1 haplotype combination of S^72^V^73^M^74^N^75^T^76^ + N^86^F^184^S^1034^N^1042^D^1246^ was most common among isolates surveyed in our study (47/122), but was seen primarily in Chennai and Nadiad and not in Rourkela. The combination of wild-type haplotypes at both loci (C^72^V^73^M^74^N^75^K^76^ + N^86^Y^184^S^1034^N^1042^D^1246^) was seen only at Rourkela, and present in the highest proportion at that site.

**Table 3:**
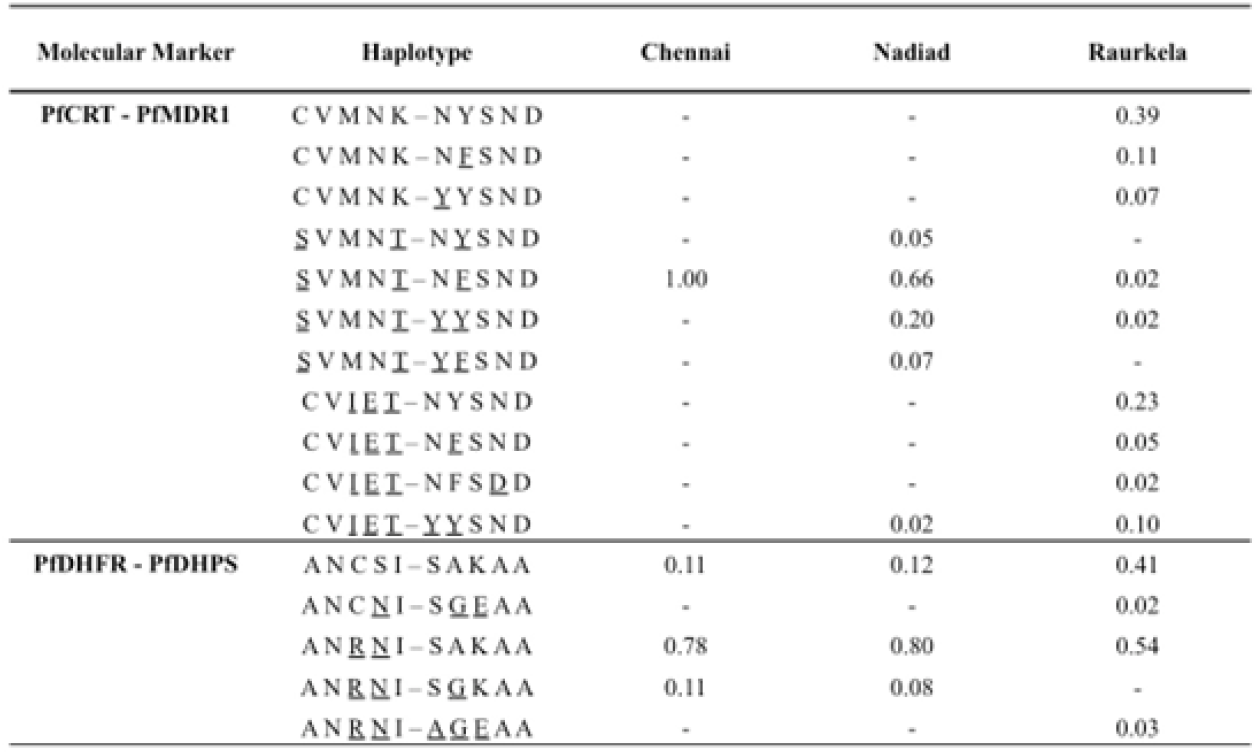
Frequency of combination haplotypes associated with drug resistance at three sites in India

### Resistance to sulfadoxine-pyrimethamine

Mutations encoding amino acid substitutions at positions 51, 59, 108, 164 in the enzyme dihydrofolate reductase PfDHFR (PF3D7_0417200) are associated with increased resistance to pyrimethamine (PYR), with the S108N mutation primarily responsible for resistance [15, 16]. 137 of 158 isolates sequenced carried only a single haplotype at the PfDHFR locus; 12 isolates carried multiple PfDHFR haplotypes and 9 isolates had partial or missing PfDHPS haplotype data (**Table 2**). The double-mutant PfDHFR haplotype A^16^N^51^R^59^N^108^I^164^ was most common among isolates (99/137) in our study, while the single-mutant A^16^N^51^C^59^ N^108^I^164^ haplotype was extremely rare (1/137); triple (A^16^I^51^R^59^N^108^I^164^) or quadruple (A^16^I^51^R^59^N^108^L^164^) mutant haplotypes could not be detected. Several isolates (37/137) had only the PYR-sensitive wild-type PfDHFR haplotype A^16^N^51^C^59^S^108^I^164^. Resistance to sulfadoxine (SUL) has been associated with mutations encoding amino acid substitutions at positions 436, 437, 540, 581 and 613 in the enzyme dihydropteroate synthetase (PfDHPS), with the A437G substitution arising first [17, 18]. 143 of 158 isolates sequenced carried only a single haplotype at the PfDHPS locus; 15 isolates had partial or missing PfDHPS haplotype data. The SUL-sensitive wild-type PfDHPS haplotype S^436^A^437^K^540^A^581^A^613^ predominated in our study, with 134/143 isolates carrying only this haplotype. Two isolates from Rourkela possessed the triple mutant PfDHPS haplotype A^436^G^437^E^540^A^581^A^613^, while the double mutant haplotype S^436^G^437^E^540^A^581^A^613^ was only detected in a single isolate. Six isolates carried the single-mutant S^436^G^437^K^540^A^581^A^613^ haplotype.

A total of 5 haplotype combinations between the PfDHFR and PfDHPS loci were seen in 129 isolates with complete sequence information for these two genes across the three sites (**Table 3**). The combination of double-mutant PfDHFR with wild-type PfDHPS (A^16^N^51^R^59^N^108^I^164^ + S^436^A^437^K^540^A^581^A^613^) was predominant among isolates in our study (87/129). Two samples from Rourkela had the strongest predictors of treatment failure in our dataset-the combination of double-mutant PfDHFR with triple-mutant PfDHPS (A^16^N^51^R^59^N^108^I^164^ + A^436^G^437^E^540^A^581^A^613^). Only 33 isolates, primarily from Rourkela, carried wild-type haplotypes at both loci. Two other haplotype combinations were observed in the population: (A^16^N^51^R^59^N^108^I^164^ + S^436^G^437^K^540^A^581^A^613^), seen in six isolates and (A^16^N^51^C^59^N^108^I^164^ + S^436^G^437^E^540^A^581^A^613^) seen in one isolate.

### Resistance to artemisinin combination therapies

Mutations in the propeller domain of the kelch protein PfK13 (PF3D7_1343700) have been shown to be associated with resistance to artemisinin derivatives [19, 20], which is defined as a delayed clearance of parasites from the blood following artemisinin administration [21]. The *Pfk13* gene was completely conserved among 92/158 isolates that were successfully sequenced at this locus, and a majority of the SNPs observed in the remaining isolates led to synonymous changes (60/92 SNPs). Only 2 isolates had substitutions in the propeller domain: V555L in N0319-V1, and A578S in R5025. We did not identify any of the four validated SNPs (C580Y, R539T, I543T or Y493H) implicated in artemisinin resistance in our isolates [19, 22]. The K189T mutation was seen in 17 isolates in our study, all from Rourkela. Mutations encoding amino acid substitutions in PfCRT, (such as I356T) are part of the genetic background associated with the emergence of artemisinin resistance [23], and were present in 20 isolates from our study (18 from Rourkela, 2 from Nadiad).

Resistance to the artemisinin combination therapy (ACT) partner drugs lumefantrine, mefloquine and piperaquine, has also been linked to certain PfCRT and PfMDR1 haplotypes. The wild type PfCRT K76 allele, associated with lumefantrine tolerance [24], was present in 46/129 isolates, all from Rourkela. The wild-type N86 allele of PfMDR1, associated with decreased susceptibility to lumefantrine [25], was observed in 103 of a total of 129 isolates in our study bearing only one PfMDR1 haplotype. The N^86^F^184^D^1246^ haplotype, selected for after artemether-lumefantrine (AL) treatment in Africa [26, 27], was observed in 58/129 isolates.

### Analysis of longitudinal Plasmodium samples from an individual

For a few enrollees (14/141), drug resistance genes were sequenced from parasite isolates collected on day 2, day 7 or day 14 following ACT treatment. Of these, three individuals were from Chennai and eleven from Nadiad. A majority of the post-treatment follow-up samples had drug resistance haplotypes that were identical to samples collected from the same patient on enrollment (**Supplementary Table 2**). However, enrollee N0319 had a mixture of wild-type (N^86^Y^184^S^1034^N^1042^D^1246^) and mutant (Y^86^F^184^S^1034^N^1042^D^1246^) PfMDR1 haplotypes on the day of enrollment, but on day 2, only the mutant (Y^86^F^184^S^1034^N^1042^D^1246^) haplotype could be detected (**Supplementary Table 2**). Most enrollees in our study did not have parasitemia detectable by microscopy or PCR on day 2 following ACT treatment.

### Using heterozygous variants for COI estimation

In order to identify possible multi-clonal infections among the clinical isolates, we ranked the isolates based on the total number of high-quality heterozygous variants (i.e. positions containing more than one allele), similar to [6] (**Figure 1**). Isolates containing more than twice the median number of heterozygous variants in the dataset were classified as potential multi-clonal infections. This included 30 isolates in total, 8 from Nadiad and 22 from Rourkela (**Supplementary Table 3**). We did not identify any multiclonal infections in Chennai.

**Figure 1.**
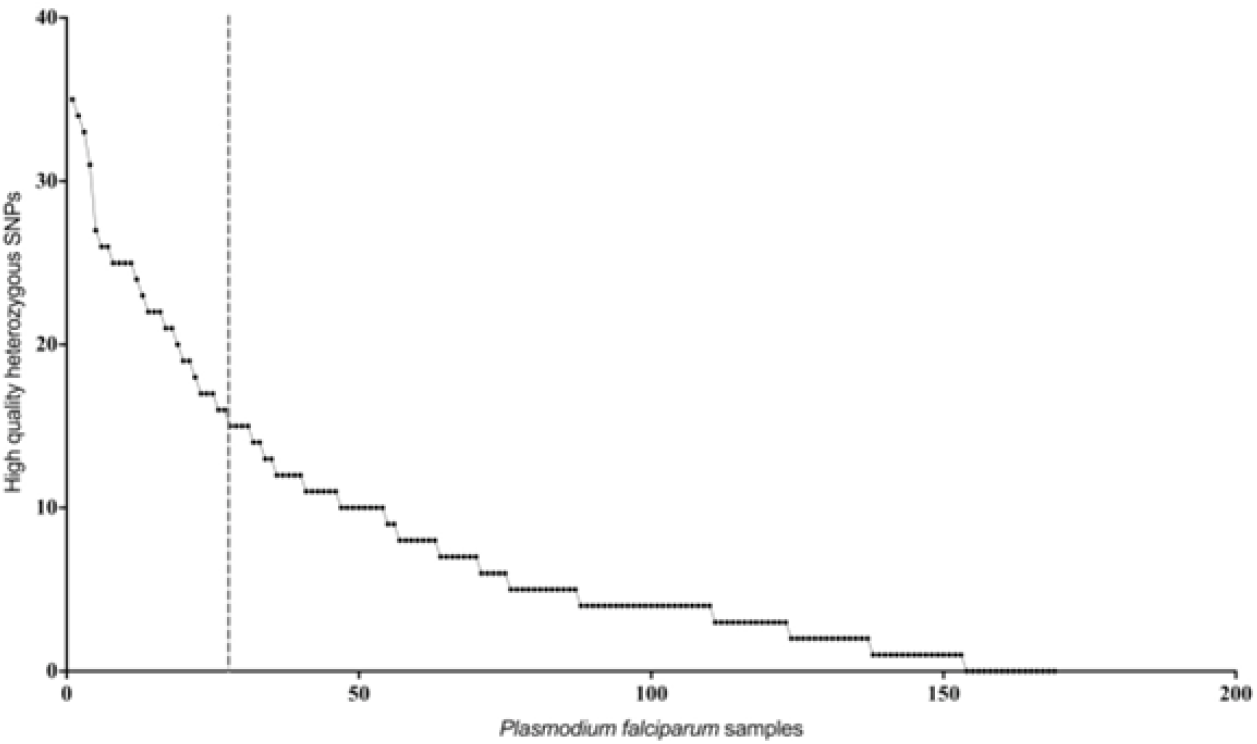
Estimation of mixed genotypes using heterozygous variant calls. Plot showing number of high quality heterozygous variant calls (y-axis) for each *P. falciparum* clinical isolate. Isolates on the left of the vertical dotted line, which represents twice the median number of heterozygous variants observed in the entire dataset, were classified as potential mixed genotype infections.

### Clustering of isolates based on drug resistance haplotypes

In order to identify population-specific variation in the drug resistance genes we performed principal component analysis (PCA) using sequence data at 230 polymorphic positions. The first two principal components accounted for 49.1% of the variation in the SNP data (PC1=30.74%, PC2=18.36%). The PCA showed isolates from Chennai and Nadiad clustering together, while isolates from Raurkela were split into two separate clusters (**Figure 2a**). Further analysis revealed that the three clusters correspond to distinct PfCRT haplotypes (codons 72-76). The two clusters of isolates from Raurkela were associated with haplotypes C^72^V^73^I^74^E^75^T^7^ and C^72^V^73^M^74^N^75^K^76^, whereas the cluster containing isolates from Chennai and Nadiad was associated with the S^72^V^73^M^74^N^75^T^76^ haplotype. Depending on the specific PfCRT haplotype, MR4 reference strains were placed across all three clusters (**Figure 2b**).

**Figure 2.**
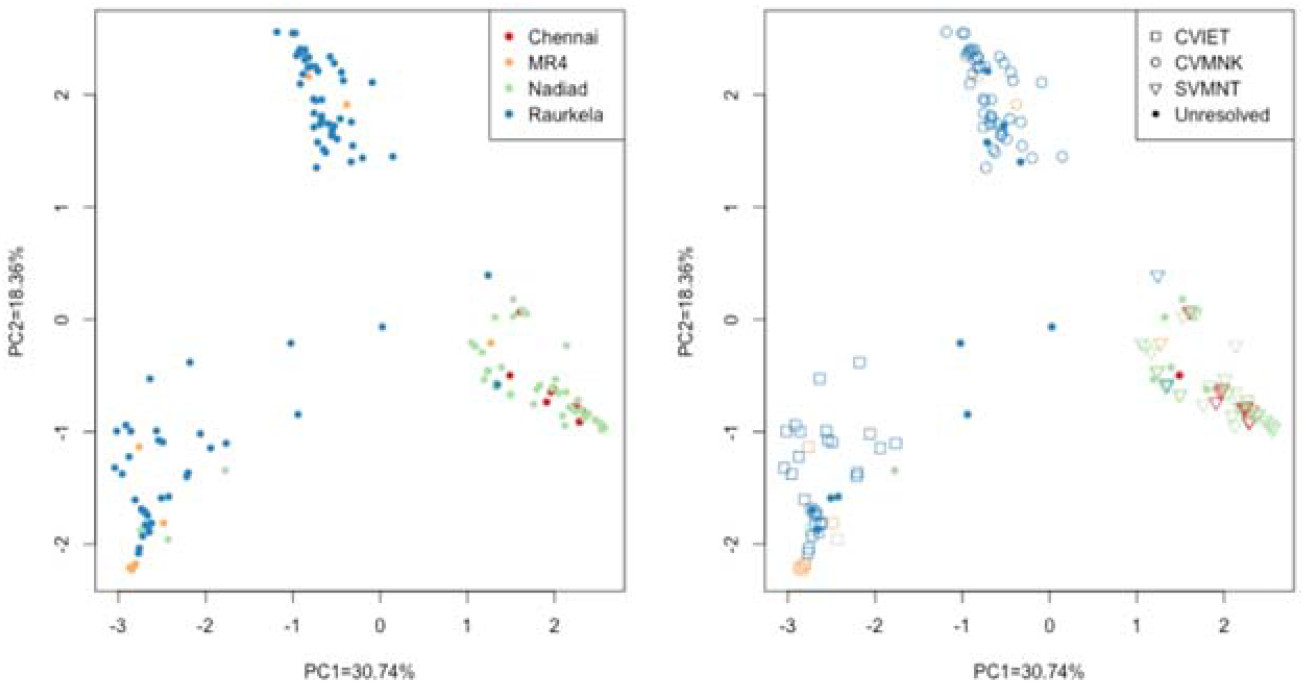
Principal component analysis using high quality SNP data from all *P. falciparum* isolates. (A) Each isolate is colored by site (B) Each isolate is colored by site and shape denotes haplotype observed at PfCRT 72-76 locus. Unresolved refers to missing or partial data and mixed infection samples.

## DISCUSSION

Tracking the emergence and spread of antimalarial drug resistance has become critical to sustaining progress towards the control and eventual elimination of malaria in South Asia, especially India. *P. falciparum* has evolved resistance to virtually every drug introduced to combat it, due to several factors such as substandard drug quality, improper prescription practices, and consumption of drugs as mono-therapy instead of combination therapies.

Chloroquine (CQ) was widely used as an antimalarial across the world since the 1930s, due to its high efficacy and relative inexpensiveness. In the 1950s CQ-resistant *P. falciparum* arose simultaneously in South America and South East Asia; it was first detected in India by 1973 and eventually spread through the country [28]. CQ remained the first-line therapy for *P. vivax* infections in India, even after its use against *P. falciparum* was discontinued. Membrane transporters such as PfCRT and PfMDR1 can acquire mutations that may modulate resistance to CQ by reducing accumulation of CQ in the food vacuole [29]. We observed a diverse distribution of three PfCRT and five PfMDR1 haplotypes in isolates across our study sites. The CQ resistance-associated PfCRT S^72^V^73^M^74^N^75^T^76^ haplotype was fixed in the Chennai and Nadiad, while parasites in Rourkela possessed either the CQ resistance-associated PfCRT C^72^V^73^I^74^E^75^T^76^ haplotype or the CQ-sensitive, wild-type PfCRT C^72^V^73^M^74^N^75^K^76^ haplotype. Among the three study sites, Rourkela has the highest prevalence of *P. falciparum*, followed by Nadiad and Chennai. Conversely, the prevalence of *P. vivax* is higher in Chennai and Nadiad, leading to higher CQ usage and drug pressure, possibly explaining the expansion of the PfCRT S^72^V^73^M^74^N^75^T^76^ haplotype at these two sites. The S^72^V^73^M^74^N^75^T^76^ haplotype from PNG [30, 31] may have been introduced to South India through Sri Lanka or the Andaman and Nicobar Islands and subsequently spread through the rest of the country [32]. It is evident through the

PCA that the parasite population in Rourkela is split into two distinct clusters, corresponding to CQ-resistant (C^72^V^73^I^74^E^75^T^76^) and CQ-sensitive (C^72^V^73^M^74^N^75^K^76^) haplotypes. While the PfCRT S^72^V^73^M^74^N^75^T^76^ haplotype is predominant in most of India, earlier studies have shown that the eastern state of Orissa has a higher prevalence of the C^72^V^73^I^74^E^75^T^76^ haplotype [33, 34]. The C^72^V^73^I^74^E^75^T^76^ haplotype may have been introduced to Rourkela through North-East India, following the well-established migratory route of South East Asian *P. falciparum* strains into India. We propose that in the absence of CQ pressure in Rourkela, there is reversion to the wild-type C^72^V^73^M^74^N^75^K^76^ haplotype from the mutant C^72^V^73^I^74^E^75^T^76^ haplotype as the fitness cost associated with the C^72^V^73^I^74^E^75^T^76^ haplotype may be greater than that of the S^72^V^73^M^74^N^75^T^76^ haplotype [35]. CQ resistance can also be modulated by mutations in PfMDR1 when they occur in the background of resistance-conferring mutations in PfCRT [35]. The PfMDR1 substitution N86Y contributes to CQ-resistance but has been associated with sensitization of *P. falciparum* parasites to dihydroartemisinin (DHA) and lumefantrine (LMF) [36]. We observed a predominance of the PfMDR1 N86 allele and PfCRT S^72^V^73^M^74^N^75^T^76^ haplotype in Chennai and Nadiad, which corroborates with previous observations in regions of low *P. falciparum* transmission in India by Mallick *et al* [37], who also identified a mixture of mutant and wild-type haplotypes in high transmission regions.

Following widespread CQ resistance, a combination of the antifolates sulfadoxine and pyrimethamine (SP) was introduced as the second-line antimalarial treatment for *P. falciparum* in 1982. Resistance to sulfadoxine (SUL) and pyrimethamine (PYR) has been associated with mutations in the folate biosynthesis pathway enzymes, *Pfdhps* and *Pfdhfr* respectively; it has been proposed that these enzymes reduce binding of the antifolates to their targets. In our study, the wild-type SUL-sensitive PfDHPS haplotype S^436^A^437^K^540^A^581^A^613^ was predominant across all three sites, occurring primarily in combination with the PYR-resistant, double-mutant PfDHFR haplotype A^16^N^51^R^59^N^108^I^164^ in Nadiad and Chennai. However, at Rourkela, there were comparable numbers of isolates with the wild type A^16^N^51^C^59^S^108^I^164^ and double-mutant A^16^N^51^R^59^N^108^I^164^ DHFR haplotypes. The double mutant A^16^N^51^R^59^N^108^I^164^ PfDHFR haplotype was predominant at each site, corroborating with previous studies from India [34, 37]. We primarily detect wild-type PfDHPS and double-mutant PfDHFR haplotypes, that confer no resistance or low-level resistance to PYR and SUL, respectively. This observation coupled with the absence of quadruple or quintuple mutant haplotypes at all three sites suggest that *P. falciparum* resistance to SP therapy at these three sites is very low [37].

Artemisinin (ART) and its derivatives-artesunate, artemether, dihydroartemisinin are among the most effective antimalarials currently available. Owing to their short half-lives in the blood, they are administered in combination with partner drugs with longer half-lives, such as lumefantrine, piperaquine, etc., in order to kill residual parasites. Artemisinin combination therapy (ACT) was recommended as first-line treatment for *P. falciparum* infections by the WHO in 2005. In most parts of India artesunate is used in combination with sulfadoxine and pyrimethamine (AS-SP), except for the North-East region where the combination of artemether and lumefantrine (AL) is used instead, due to widespread SP resistance [9]. Artemisinin resistance was first identified in Cambodia and spread quickly through South East Asia, while also emerging simultaneously at different foci in South Asia. Mutations in the propeller domain of the kelch protein K13 were shown to be associated with delayed clearance of parasites after ACT treatment [19, 20]. Since K13 mutations were recently detected in an area of Myanmar that is adjacent to the eastern border of India [38], there has been concern about the spread of artemisinin resistance into India, in a manner similar to the previous spread of CQ and SP resistance. If artemisinin fails, there is no drug with comparable efficacy to replace it. While ACT failure has been reported in North East India, it may be attributed to failure of the partner drugs [39, 40], and studies examining the propeller domain have not detected any mutations associated with resistance among Indian isolates except F446I, which remains to be validated [41-43]. Our study identified two mutations in the propeller region – one from an isolate in Nadiad, and the other from Rourkela. The V555L substitution (seen in sample N0319-V1) has been detected in Senegal at a very low frequency [44], while A578S (seen in R5025) has been detected in several isolates from Africa, India and Bangladesh [22, 41, 42, 45]. Neither has been associated with resistance among field isolates or in *in vitro* assays. The K189T mutation that was identified in a subset of the Rourkela population has previously been seen in isolates from Senegal, Guyana, the China-Myanmar border, and Northeast India [42, 46-48]. The wild-type N86 allele of PfMDR1 and the wild-type K76 allele of PfCRT were selected after artemether-lumefantrine combination therapy in Africa [24-27] and were associated with decreased susceptibility to lumefantrine *in vitro* [24]. Our data suggests that AL treatment is likely be effective in areas with a predominance of CQ-resistant PfCRT haplotypes S^72^V^73^M^74^N^75^T^76^ (Chennai and Nadiad) and C^72^V^73^I^74^E^75^T^76^ (Rourkela) as well as the PfMDR1 haplotypes Y^86^F^184^S^1034^N^1042^D^1246^ and Y^86^Y^184^S^1034^N^1042^D^1246^ (Nadiad and Rourkela).

Parasite isolates from Rourkela showed greater diversity than isolates from Chennai and Nadiad at all drug resistance loci. PCA clustering revealed that parasites from Chennai and Nadiad were more closely related, whereas the parasites in Rourkela separated into two populations that were associated with distinct PfCRT haplotypes. There was a near-equal distribution of wild-type and mutant haplotypes at the Pfcrt, Pfdhfr and Pfmdr1 loci in Rourkela. The eight isolates in our dataset that possessed wild-type haplotypes at all the drug resistance loci were also from Rourkela. Interestingly, over 70% of the potential multi-clonal infections identified were from Rourkela, the remainder from Nadiad, and none were from Chennai. We suspect that as a result of higher prevalence and transmission of P. falciparum in Rourkela, the complexity of infection may be higher at this site, compared to Nadiad or Chennai, where the prevalence of P. falciparum is much lower.

We have demonstrated the potential of amplicon sequencing to discriminate between single- and multi-clonal infections. Another application of this method is to study the variation in within-host diversity in response to chemotherapy. This is particularly interesting for multi-clonal infections as it can enable us to track changes in the frequency of individual alleles before and after drug treatment [49]. Our analysis of parasite isolates from follow-up (collected after ACT-treatment) samples was limited to a small number of samples from Chennai and Nadiad. For all pairs of isolates collected from same individual, the key drug resistance SNPs were identical enrollment and follow-up isolates, except in the case of one patient isolate in Nadiad (N0319) that possessed a mixture of CQ-resistant and CQ-sensitive haplotypes at the *Pfmdr1* locus at enrollment, but retained only the CQ-resistant haplotype following drug treatment. The potential multi-clonal infections identified in our study did not include any pairs of follow-up isolates, mainly because we did not have any follow-up samples collected in Rourkela. The likelihood of the pretreatment infection being multi-clonal is much lower in Chennai and Nadiad, compared to Rourkela because of lower COI. Therefore, it is not surprising that we identified the same parasite haplotypes in patients, pre-and post-ACT administration.

We have established amplicon sequencing as an important tool for high-throughput molecular surveillance of antimalarial drug resistance in India, especially since it can be implemented in field settings, as we have demonstrated in this study.

## Data Availability

All data produced in the present study are available upon reasonable request to the authors

## Abbreviations

**MSP-1**_**19**_: merozoite surface protein-1_19_

**AMA-1:** apical membrane antigen-1

## Declarations

### Ethics approval and consent to participate

Ethical approval to conduct this study was obtained from New York University Institutional Review Board (Study #i10-00173) and the Ethics Committee of the National Institute of Malaria Research, India (ICMR). All project staff completed Protection of Human Research Subjects training prior to beginning the study, and clinical samples were collected after informed consent was obtained from all participants.

## Author’s Contributions

JMC, OPS, PKM, designed the study. OPS, JMC, PKM contributed reagents/materials/analysis tools. AA, SR, SLPG, NT, SZA, HCS, SKS, AE contributed in sample collection and processing. SK, NB performed the experiments. SK, SU, PR, PKM analyzed and interpreted data. SK, SU, PR wrote the first draft. PKM, OPS, JMC reviewed and edited the manuscript. All authors read and approved the final manuscript.

## Acknowledgments

We thank MR4 for providing malaria parasite strains. We are indebted to Drs. Lalitha Ramanathapuram, Patrick Sutton, Anna Maria van Eijk and all members of the CSCMi teams in Delhi, Nadiad, Raurkela and Chennai for their support. Dr. Martina Bradic’s help with the linkage analysis is greatly appreciated. We also thank Dr. Neena Valecha and the staff of the National institute of Malaria Research in Delhi, India for use of the facilities.

## Supplementary Excel File

Supplementary Table 1: List of annotated, high-quality SNPs in six P. falciparum genes reported by the Ion Torrent PGM Variant Caller Plugin

Supplementary Table 2: Data for follow-up patient samples showing key SNPs implicated in drug resistance from six P. falciparum genes (represented as haplotypes).

Supplementary Table 3: High-quality SNPs in samples classified as potential multi-clonal infections

## REFERENCES

1. Wangdi, K., et al., Malaria elimination in India and regional implications. Lancet Infect Dis, 2016. 16(10): p. e214–24.

2. WHO, World Malaria Report 2016. 2016.

3. WHO, World Malaria Report 2015-December 2015. 2015.

4. NVBDCP, National Framework for Malaria Elimination in India 2016–2030

5. Shah, N.K., et al., Antimalarial drug resistance of Plasmodium falciparum in India: changes over time and space. Lancet Infect Dis, 2011. 11(1): p. 57–64.

6. Rao, P.N., et al., A Method for Amplicon Deep Sequencing of Drug Resistance Genes in Plasmodium falciparum Clinical Isolates from India. J Clin Microbiol, 2016. 54(6): p. 1500–11.

7. Das, A., et al., Malaria in India: the center for the study of complex malaria in India. Acta Trop, 2012. 121(3): p. 267–73.

8. National Health Mission Tamil Nadu, D.o.H.a.F.W.; Available from: http://www.nrhmtn.gov.in/vbdc.html.

9. Anvikar, A.R., et al., Antimalarial drug policy in India: Past, present & future, in Indian J Med Res. 2014. p. 205–15.

10. Sidhu, A.B., D. Verdier-Pinard, and D.A. Fidock, Chloroquine resistance in Plasmodium falciparum malaria parasites conferred by pfcrt mutations. Science, 2002. 298(5591): p. 210–3.

11. Fidock, D.A., et al., Mutations in the P. falciparum digestive vacuole transmembrane protein PfCRT and evidence for their role in chloroquine resistance. Mol Cell, 2000. 6(4): p. 861–71.

12. Djimde, A., et al., Application of a molecular marker for surveillance of chloroquine-resistant falciparum malaria. Lancet, 2001. 358(9285): p. 890–1.

13. Vathsala, P.G., et al., Widespread occurrence of the Plasmodium falciparum chloroquine resistance transporter (Pfcrt) gene haplotype SVMNT in P. falciparum malaria in India. Am J Trop Med Hyg, 2004. 70(3): p. 256–9.

14. Menard, D. and A. Dondorp, Antimalarial Drug Resistance: A Threat to Malaria Elimination. Cold Spring Harb Perspect Med, 2017.

15. Cowman, A.F., et al., Amino acid changes linked to pyrimethamine resistance in the dihydrofolate reductase-thymidylate synthase gene of Plasmodium falciparum. Proc Natl Acad Sci U S A, 1988. 85(23): p. 9109–13.

16. Sirawaraporn, W., et al., Antifolate-resistant mutants of Plasmodium falciparum dihydrofolate reductase. Proc Natl Acad Sci U S A, 1997. 94(4): p. 1124–9.

17. Triglia, T., et al., Allelic exchange at the endogenous genomic locus in Plasmodium falciparum proves the role of dihydropteroate synthase in sulfadoxine-resistant malaria. EMBO J, 1998. 17(14): p. 3807–15.

18. Brooks, D.R., et al., Sequence variation of the hydroxymethyldihydropterin pyrophosphokinase: dihydropteroate synthase gene in lines of the human malaria parasite, Plasmodium falciparum, with differing resistance to sulfadoxine. Eur J Biochem, 1994. 224(2): p. 397–405.

19. Straimer, J., et al., Drug resistance. K13-propeller mutations confer artemisinin resistance in Plasmodium falciparum clinical isolates. Science, 2015. 347(6220): p. 428–31.

20. Ariey, F., et al., A molecular marker of artemisinin-resistant Plasmodium falciparum malaria. Nature, 2014. 505(7481): p. 50–5.

21. Dondorp, A.M., et al., Artemisinin resistance in Plasmodium falciparum malaria. N Engl J Med, 2009. 361(5): p. 455–67.

22. Menard, D., et al., A Worldwide Map of Plasmodium falciparum K13-Propeller Polymorphisms. N Engl J Med, 2016. 374(25): p. 2453–64.

23. Miotto, O., et al., Genetic architecture of artemisinin-resistant Plasmodium falciparum. Nat Genet, 2015. 47(3): p. 226–34.

24. Sisowath, C., et al., In vivo selection of Plasmodium falciparum parasites carrying the chloroquine-susceptible pfcrt K76 allele after treatment with artemether-lumefantrine in Africa. J Infect Dis, 2009. 199(5): p. 750–7.

25. Sisowath, C., et al., In vivo selection of Plasmodium falciparum pfmdr1 86N coding alleles by artemether-lumefantrine (Coartem). J Infect Dis, 2005. 191(6): p. 1014–7.

26. Dokomajilar, C., et al., Selection of Plasmodium falciparum pfmdr1 alleles following therapy with artemether-lumefantrine in an area of Uganda where malaria is highly endemic. Antimicrob Agents Chemother, 2006. 50(5): p. 1893–5.

27. Humphreys, G.S., et al., Amodiaquine and artemether-lumefantrine select distinct alleles of the Plasmodium falciparum mdr1 gene in Tanzanian children treated for uncomplicated malaria. Antimicrob Agents Chemother, 2007. 51(3): p. 991–7.

28. Sehgal, P.N.S. M. I. D.; Sharma, S. L.; Gogai, S., Resistance to chloroquine in falciparum malaria in Assam State, India‥ Journal of Communicable Diseases, 1973. 5(4): p. 175–180.

29. Saliba, K.J., P.I. Folb, and P.J. Smith, Role for the plasmodium falciparum digestive vacuole in chloroquine resistance. Biochem Pharmacol, 1998. 56(3): p. 313–20.

30. Mehlotra, R.K., et al., Evolution of a unique Plasmodium falciparum chloroquine-resistance phenotype in association with pfcrt polymorphism in Papua New Guinea and South America. Proc Natl Acad Sci U S A, 2001. 98(22): p. 12689–94.

31. Mixson-Hayden, T., et al., Evidence of selective sweeps in genes conferring resistance to chloroquine and pyrimethamine in Plasmodium falciparum isolates in India. Antimicrob Agents Chemother, 2010. 54(3): p. 997–1006.

32. Mallick, P.K., et al., Microsatellite analysis of chloroquine resistance associated alleles and neutral loci reveal genetic structure of Indian Plasmodium falciparum. Infect Genet Evol, 2013. 19: p. 164–75.

33. Sutar, S.K., et al., Sequence analysis of coding DNA fragments of pfcrt and pfmdr-1 genes in Plasmodium falciparum isolates from Odisha, India. Mem Inst Oswaldo Cruz, 2011. 106(1): p. 78–84.

34. Kar, N.P., et al., Comparative assessment on the prevalence of mutations in the Plasmodium falciparum drug-resistant genes in two different ecotypes of Odisha state, India. Infect Genet Evol, 2016. 41: p. 47–55.

35. Sa, J.M., et al., Geographic patterns of Plasmodium falciparum drug resistance distinguished by differential responses to amodiaquine and chloroquine. Proc Natl Acad Sci U S A, 2009. 106(45): p. 18883–9.

36. Veiga, M.I., et al., Globally prevalent PfMDR1 mutations modulate Plasmodium falciparum susceptibility to artemisinin-based combination therapies. Nat Commun, 2016. 7: p. 11553.

37. Ahmed, A., et al., Plasmodium falciparum isolates in India exhibit a progressive increase in mutations associated with sulfadoxine-pyrimethamine resistance. Antimicrob Agents Chemother, 2004. 48(3): p. 879–89.

38. Tun, K.M., et al., Spread of artemisinin-resistant Plasmodium falciparum in Myanmar: a cross-sectional survey of the K13 molecular marker. Lancet Infect Dis, 2015. 15(4): p. 415–21.

39. Saha, P., et al., Comparative efficacies of artemisinin combination therapies in Plasmodium falciparum malaria and polymorphism of pfATPase6, pfcrt, pfdhfr, and pfdhps genes in tea gardens of Jalpaiguri District, India. Antimicrob Agents Chemother, 2012. 56(5): p. 2511–7.

40. Mishra, N., et al., Declining efficacy of artesunate plus sulphadoxine-pyrimethamine in northeastern India. Malar J, 2014. 13: p. 284.

41. Mishra, N., et al., Surveillance of artemisinin resistance in Plasmodium falciparum in India using the kelch13 molecular marker. Antimicrob Agents Chemother, 2015. 59(5): p. 2548–53.

42. Mishra, N., et al., Emerging polymorphisms in falciparum Kelch 13 gene in Northeastern region of India. Malar J, 2016. 15(1): p. 583.

43. Chatterjee, M., et al., No Polymorphism in Plasmodium falciparum K13 Propeller Gene in Clinical Isolates from Kolkata, India. J Pathog, 2015. 2015: p. 374354.

44. Talundzic, E., et al., Molecular Epidemiology of Plasmodium falciparum kelch13 Mutations in Senegal Determined by Using Targeted Amplicon Deep Sequencing. Antimicrob Agents Chemother, 2017. 61(3).

45. Mohon, A.N., et al., Mutations in Plasmodium falciparum K13 propeller gene from Bangladesh (2009-2013). Malar J, 2014. 13: p. 431.

46. Torrentino-Madamet, M., et al., Limited polymorphisms in k13 gene in Plasmodium falciparum isolates from Dakar, Senegal in 2012-2013. Malar J, 2014. 13: p. 472.

47. Wang, Z., et al., Prevalence of K13-propeller polymorphisms in Plasmodium falciparum from China-Myanmar border in 2007-2012. Malar J, 2015. 14: p. 168.

48. Rahman, R., et al., Continued Sensitivity of Plasmodium falciparum to Artemisinin in Guyana, With Absence of Kelch Propeller Domain Mutant Alleles. Open Forum Infect Dis, 2016. 3(3): p. ofw185.

49. Mideo, N., et al., Ahead of the curve: next generation estimators of drug resistance in malaria infections. Trends Parasitol, 2013. 29(7): p. 321–8.

